# Gaps in mobility data and implications for modelling epidemic spread: a scoping review and simulation study

**DOI:** 10.1101/2022.03.07.22272001

**Authors:** Jack Wardle, Sangeeta Bhatia, Moritz U G Kraemer, Pierre Nouvellet, Anne Cori

**Affiliations:** MRC Centre for Global Infectious Disease Analysis, School of Public Health, Imperial College London, UK; Department of Zoology, University of Oxford, Oxford, UK; School of Life Sciences, University of Sussex, Brighton, UK

## Abstract

Reliable estimates of human mobility are important for understanding the spatial spread of infectious diseases and the effective targeting of control measures. However, when modelling infectious disease dynamics, data on human mobility at an appropriate temporal or spatial resolution are not always available, leading to the common use of model-derived mobility proxies. In this study we reviewed the different data sources and mobility models that have been used to characterise human movement in Africa. We then conducted a simulation study to better understand the implications of using human mobility proxies when predicting the spatial spread and dynamics of infectious diseases.

We found major gaps in the availability of empirical measures of human mobility in Africa, leading to mobility proxies being used in place of data. Empirical data on subnational mobility were only available for 17/54 countries, and in most instances, these data characterised long-term movement patterns, which were unsuitable for modelling the spread of pathogens with short generation times (time between infection of a case and their infector). Results from our simulation study demonstrated that using mobility proxies can have a substantial impact on the predicted epidemic dynamics, with complex and non-intuitive biases. In particular, the predicted times and order of epidemic invasion, and the time of epidemic peak in different locations can be underestimated or overestimated, depending on the types of proxies used and the country of interest.

Our work underscores the need for regularly updated empirical measures of population movement within and between countries to aid the prevention and control of infectious disease outbreaks. At the same time, there is a need to establish an evidence base to help understand which types of mobility data are most appropriate for describing the spread of emerging infectious diseases in different settings.

## Introduction

Human movement is a key determinant of the spatial spread of infectious diseases as evidenced by the spread of Ebola [1, 2], yellow fever [3, 4], and most recently, SARS-CoV-2 [5, 6]. The emergence and subsequent international spread of the latter and its variants of concern (VOCs) highlight the speed and scale at which infectious pathogens can spread around the globe [7–9].

Models of infectious disease dynamics that incorporate human movement have been used to generate insights into the spatial spread of pathogens. They can help identify locations that are most susceptible to receive imported cases [10–12], assess the effectiveness of travel restrictions [13, 14], and inform the allocation of scarce resources such as vaccines, drugs, personal protective equipment or specialist healthcare staff [15–18]. Spatial models of infectious disease spread typically rely on human population movement data. These can be empirical measurements, for example flight passenger statistics, information from national population censuses, or mobile phone call detail records [19, 20]. More recently, a few high resolution datasets based on mobile phone GPS data have been made available to researchers and public health officials and used to analyse within-country movement patterns during the COVID-19 pandemic [21–23].

However, empirical data on human movement are not routinely collected, particularly in low- and middle-income countries [24]. Moreover, where empirical data are available, these are not always at a spatial and temporal resolution that matches the scale and pace at which pathogens spread [25]. Although mobile phone usage is increasingly being used to understand movement of populations at a fine resolution, these data are known to be biased in systematic ways by limited smartphone ownership or uneven mobile phone coverage [26, 27], and typically do not capture international movement. They are also expensive and time-consuming to acquire, limiting their potential use in real-time analysis. Finally, privacy concerns constrain wider sharing of these data [28], preventing the reproducibility of analyses based on them.

In the absence of empirical data, infectious disease modelling studies often rely on human movement models such as gravity and radiation models [29–31]. These models are sometimes calibrated using data from nearby countries [4, 29], long-term movement data from Demographic and Health Surveys (DHS), or observed patterns of disease spread [32]. By definition, these mobility proxies cannot be validated against local human movement data which do not exist. The potential impact of using such proxies to predict the spatial dynamics of infectious diseases has received little attention despite their recurring role in informing policy and resource allocation decisions.

Therefore, better understanding both the availability of human mobility data, or lack thereof, and the impact of using mobility proxies on predicting infectious disease spread is key to prioritise future research effort. Here we focus on the African continent which over the past decade has experienced numerous large epidemics with significant public health consequences, including multiple epidemics of Ebola [33], yellow fever [34], cholera [35], and measles [36]. We first reviewed the different data sources and mobility models that have been used to estimate human movement in Africa. We then conducted a simulation study, based on high resolution movement data from Europe, to better understand the implications of using human mobility proxies when predicting the spatial spread of infectious diseases.

## Methods

### Search Strategy

We carried out a scoping review of mobility data and models in Africa, adhering to the guidelines established by PRISMA extension for Scoping Reviews [37]. We searched PubMED and Web of Science on 29^th^ August 2018 for peer reviewed literature in English on human movement data used in mobility models in Africa. We restricted our search to gravity and radiation models, which are the most commonly used in infectious disease modelling [38–42].

In short, in a gravity model the population movement between two locations is assumed to be proportional to the population sizes of the origin and distance and inversely proportional to the distance between them (Eq. (3)). The radiation model also describes how the movement between two locations may be affected by other surrounding highly populated locations (Eq. (S1)).

The search terms used were: (gravity model OR radiation model) AND human AND africa), (mobility model AND africa AND human), (gravity model AND africa), (radiation model AND africa AND human). Both abstract and full-text screening were carried out independently by two reviewers and all disagreements were resolved by consensus. Only primary research articles were eligible for inclusion. We excluded studies that were not set in Africa, did not have a spatial component, or did not use data or estimates of human mobility.

Data were systematically extracted from the studies to describe the source (e.g. census), spatial resolution, and temporal resolution of data on human movement. We extracted the location and the time period of the data for each study and whether the data were made available.

We classified as “empirical” human movement data obtained from mobile phone call detail records (CDR), micro-census data from Integrated Public Use Microdata Series (IPUMS), short-term migration data from DHS, and data on long-term migration (Global Bi-lateral Migration Database, hereafter GBMD), or on movement of refugees from the United Nations High Commissioner for Refugees (UNHCR). In addition to these generic data sources, some studies carried out surveys tailored to identify movement patterns relevant to the spread of specific diseases, e.g. overnight stays for malaria. These were also considered as empirical data sources. In contrast to these, a large number of studies used models of human mobility calibrated to data from nearby regions, or to the observed patterns of disease spread. Some studies specifically aimed to generate such mobility estimates for regions where no empirical data were available. Where a study did not rely on empirical measurements to characterise mobility and instead used indirect sources such as disease spread or data from other countries, we refer to it as having used (or produced) “mobility proxies”. For those studies, we also extracted details of the mobility models used. For each country included in a study, we considered empirical data (or mobility proxy) for a given spatial resolution and for a specific time period as a discrete data set.

### Simulation Study

To understand the implications of using human mobility proxies instead of empirical data when predicting the spatial spread and dynamics of infectious diseases, we carried out a simulation study comparing epidemic outcomes when using empirical movement data versus mobility proxies.

### Commuting data

The settings for our simulation study were chosen based on locations where empirical mobility data were openly accessible and therefore were not set in Africa. We used commuting patterns derived by Tizzoni <u>et al</u>. from mobile phone call data in France and Portugal for 2007 and 2006 respectively [43]. The commuting patterns were obtained from a large sample of users’ CDR by identifying the frequently visited locations (based on proximity to mobile phone towers) of each person in the dataset. The authors assumed that the most visited place corresponded to the location where an individual lives, with the second most visited place being the location where an individual works [44]. This was then used to calculate the total numbers of people in the sample who commute between each of the 323 districts (corresponding to the ADM3 administrative units) in mainland France and the 278 municipalities (the ADM2 administrative units) in mainland Portugal. This approach also gave the number of people that both lived and worked in the same spatial unit.

We combined the commutes between and within administrative units to construct an origin-destination (O-D) movement matrix for each country. To generate population-level estimates of the total movement, we scaled the sampled movement flows between pairs of administrative units. The total number of individuals living in spatial unit *i* and working in unit *j* is calculated as:

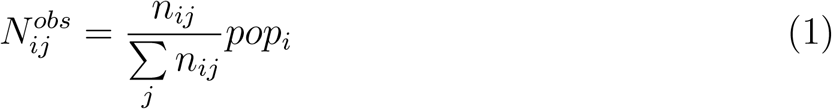

where *n*_*ij*_ is the number of people commuting from *i* to *j* in the sample of mobile phone users, Σ_*j*_ *n*_*ij*_ is the total number of people living in *i* that were included in the mobile phone data sample and *pop*_*i*_ is the population of location *i*. The scaling method in Eq. (1) ensures that the sum of each row in the O-D matrix equals the population living in that administrative unit 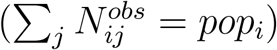.

### Mobility proxies

In order to assess the impact of using mobility proxies in epidemic models, we constructed predicted movement matrices at the same spatial resolution as the observed data in France and Portugal. Movement matrices can encode either the total number of individuals (referred to as O-D matrix) or the probability of moving between locations (rescaled O-D matrix). For the latter, matrix entry *p*_*ij*_ is the probability that an individual moves between spatial units *i* and *j*. This is the product of the probability that i) they leave *i* for any destination 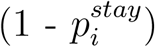; and ii) they move to *j* given that they have moved out of *i* (for which we use the notation 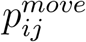). This overall probability can be written as:

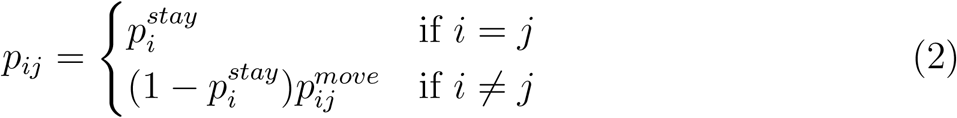

where 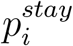 is the probability that a person living in *i* also works in that same location.

Predictions of the relative flows 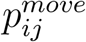 were obtained from the gravity model, which posits that the flow of individuals from location *i* to location *j* is proportional to:

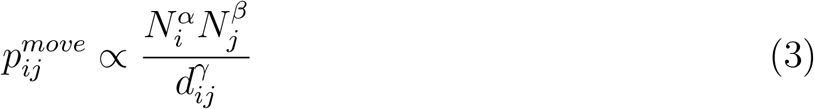

where *N*_*i*_ and *N*_*j*_ are the populations living in locations *i* and *j*, and *d*_*ij*_ is the distance between the two locations [30, 45]. *α, β* and *γ* are model parameters. We fitted the gravity model to observed movement data and then used the estimated parameters to predict 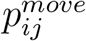.

The gravity model in Eq. (3) cannot be used to estimate 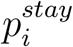. Therefore, 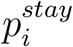 values were taken directly from observed data and used to populate the diagonal elements of the *p*_*ij*_ matrix and to scale the off-diagonal elements as shown in Eq. (2).

We multiplied the rescaled O-D matrix by the size of the origin population (*pop*_*i*_) to obtain the numbers of people predicted to move between pairs of spatial units (O-D matrix):

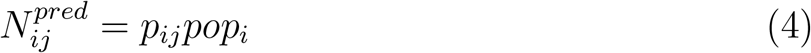

We generated different mobility proxies 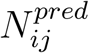 by varying assumptions about the availability of movement data in a country and about 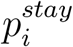. In our central scenario presented here, we assume that movement data are unavailable for a given country and we therefore make predictions based on data from a nearby country (Fig. 1). For Portugal, we generate mobility proxies using data from France, i.e. 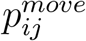 estimated by a gravity model fitted to France and 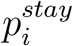 set to the average observed 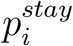 across all French administrative units. The same approach was used to generate mobility proxies for France using data from Portugal.

**Figure 1:**
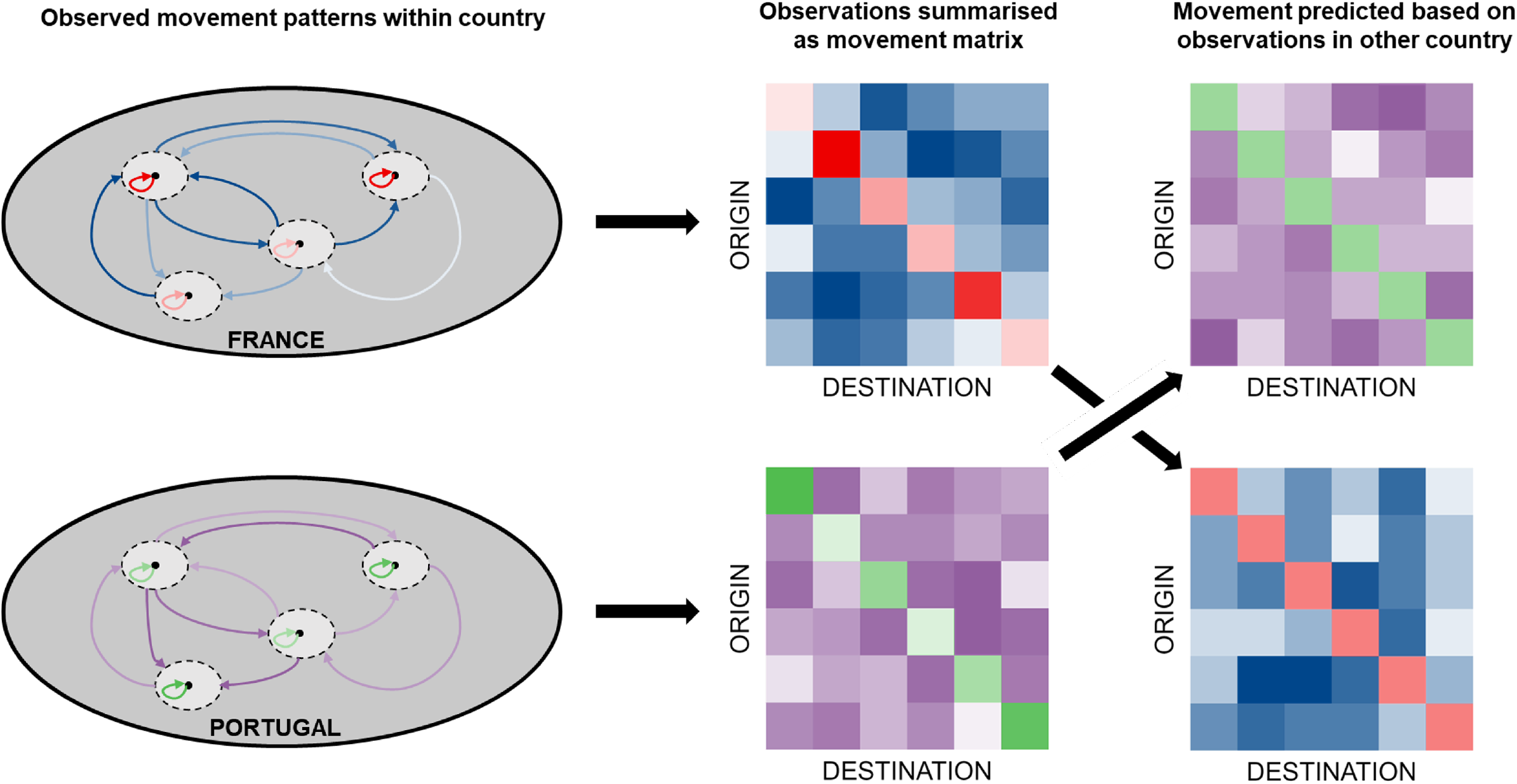
Illustration of how movement patterns observed in France and Portugal can be used to generate mobility proxies. The ovals on the left represent the movement patterns within each country, with the smaller dashed ovals representing different spatial units within the country. Movement between locations is shown by the blue arrows in France and purple arrows in Portugal. Darker colours indicate more movement. People that move within their own spatial unit are shown with red arrows in France and green arrows in Portugal. These movement patterns can be summarised in a movement matrix (i.e. O-D matrices, middle column) that provide information on movements between each origin-destination pair (including where the destination is the same as the origin, i.e. 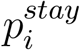, see methods). The matrices in the right column illustrate how movement patterns in France are predicted in our central scenario based on a combination of a gravity model fitted to Portuguese movement data along with data on the probability that an individual leaves the location they live in (1 - 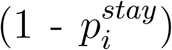. Similarly, we predict Portuguese movement using a gravity model fitted to French movement data and the average 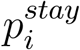 observed in France.

We explored additional scenarios where 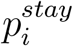 and 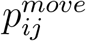 were informed by data from either the correct or the neighbouring country and at various spatial resolutions (e.g. gravity model fitted to ADM2 but used to predict movement at ADM3 in the same country) (Suppl Tab. S1). To explore the robustness of our results to the choice of mobility model, we also considered a scenario using a radiation model to predict 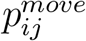.

### Epidemic model

We use a stochastic discrete time SEIR metapopulation model to simulate epidemics in France and Portugal. The subpopulations in the metapopulation model are formed from each combination of home and work location. The size of each subpopulation *N*_*ij*_ is the number of individuals who live in *i* and work in *j. N*_*ij*_ is set using either the observed data 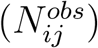 or mobility proxy 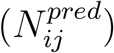. This means that the home and work locations of individuals are fixed within our model, with movements occurring recurrently. People in each subpopulation fall into one of four compartments denoting their infection states: susceptible (*S*), exposed (*E*), infectious (*I*) or recovered (*R*).

Each subpopulation has two sets of interactions with other subpopulations. For half of each day (a day being defined as a 24-hour period), we assume homogeneous mixing between all individuals who reside in the same spatial unit (i.e. all people living in *i*) and for the other half of the day we assume homogeneous mixing between individuals who work in the same spatial unit (all people working in *j*). These assumptions are in line with previous studies that incorporated commuting data in models of infectious disease spread [43, 46].

Thus there are two forces of infection acting each day on every subpopulation in our model:

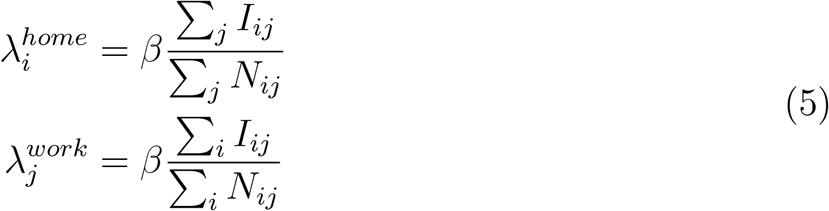

where *β* is the per capita transmission rate. 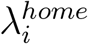 therefore depends on the number of infectious people living in the same spatial unit *i* (Σ_*j*_ *I*_*ij*_). Similarly, 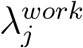 depends on the number of infectious people working in the same spatial unit *j* (Σ_*i*_ *I*_*ij*_). We make the simplifying assumption that movement is independent of infection state and that *β* is the same in all settings.

The transitions between infection compartments at each time step (Δ*t* = 0.1 day) are modelled stochastically as follows:

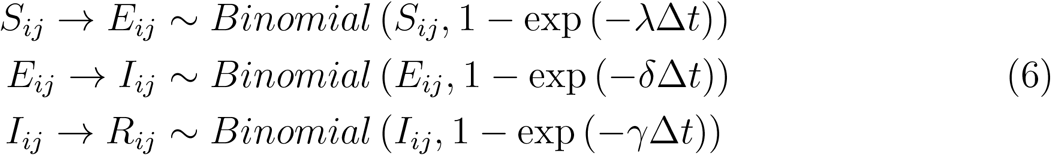

where 1*/δ* and 1*/γ* are the mean latent and infectious periods respectively. *λ* takes the value 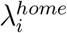 in the first half of each day when people are at home, and 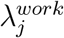 in the second half of each day when people are at work.

We simulated and compared epidemics in France and Portugal under different scenarios, where *N*_*ij*_ was informed by different data. In the baseline scenario, *N*_*ij*_ was set to observed data 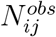, while in the alternative scenarios *N*_*ij*_ was based on mobility proxies 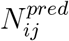 (Suppl Tab. S1). The epidemics were seeded in either the capital city (Paris and Lisbon) or a less central location (Brest in the northwest of France and Miranda do Douro in the northeast of Portugal). These locations were chosen to explore differences in the dynamics of outbreaks originating in a prominent urban centre and a location on the edge of the commuting network.

For each scenario, we ran 100 simulated epidemics for 300 days, seeding 5 infectious cases of a flu-like pathogen and assuming that the rest of the population is fully susceptible (see Suppl Sec. 2 for further details). We restricted our analysis to epidemics which were seeded successfully (defined as simulations where *>* 10% of the total population was infected after 300 days).

For each scenario *s*, we summarised all the successfully seeded epidemic simulations with four metrics, two which characterise scenario *s* only, and two which compare *s* to the baseline:

- **Invasion time in** 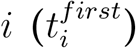, defined as the median time to the arrival of the first infectious case among those living in location *i*.
- **Peak time in** 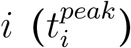, defined as the median time to the epidemic peak in location *i* (i.e. time when the number of infectious individuals living in *i* is at a maximum).
- **Relative error in invasion time in** 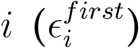. This was calculated as the difference between 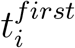 in scenario *s* (i.e. using mobility proxy) and in the baseline (using the empirical mobility data), divided by the largest invasion times across all locations in scenario *s*: 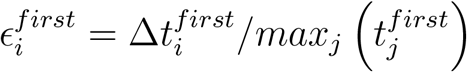.
- **Invasion order similarity** 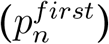, defined as the proportion of the first *n* locations invaded in the baseline scenario that were also among the first *n* locations invaded in scenario *s*. The invasion order was defined based on the median invasion time across all simulations with successfully seeded epidemics, i.e. 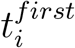.

## Results

### Scoping Review

Of the 471 articles from the initial search, 129 were selected for abstract screening and 30 full-text articles for data extraction, all of which were published between 2007 and 2018. Across the 30 studies, we identified 150 empirical human mobility data sets and 168 mobility proxies.

### Empirical data

Empirical data were available for 52 of 54 African countries (Fig. 2). For 51 countries, data on long-term migration patterns between countries were available, from GBMD (n = 42 countries) or data on refugee movement from UNHCR (n = 51). The GBMD provides data on international migrations between all countries for each decade in the period 1960–2000 [47]. The UNHCR collects data on the annual flows of refugees, asylum seekers and the number of internally displaced persons between a country/territory of origin and asylum [48].

**Figure 2:**
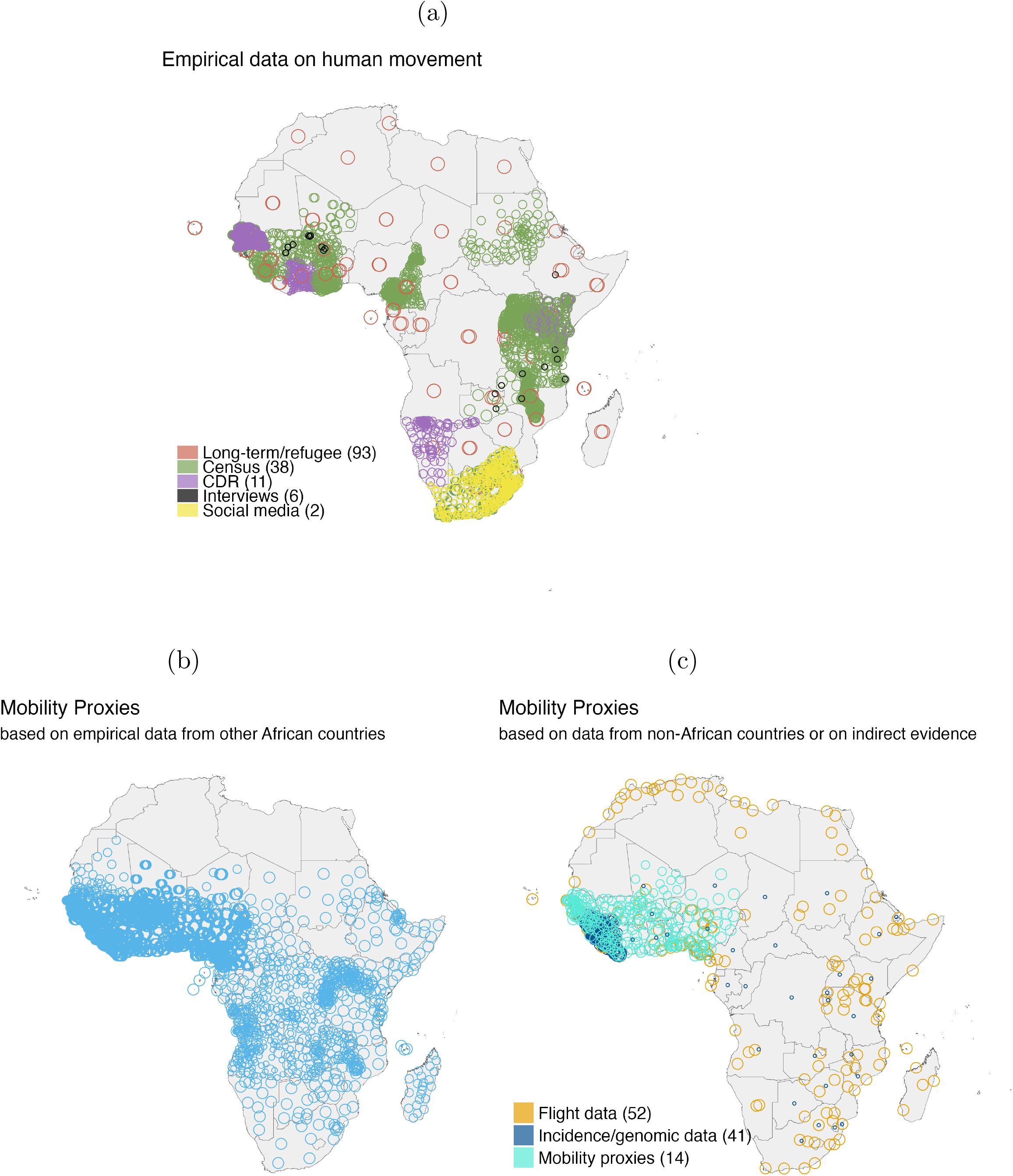
Human population mobility data and proxies in Africa. We reviewed 30 peer-reviewed articles using human mobility data or proxies in any African country. For each country included in a study, we considered empirical data (or mobility proxy) for a given spatial resolution and for a specific time period as a discrete data set. We identified 150 empirical data sets and 168 mobility proxies. (a) Locations for which empirical data on human mobility were available. Data from Global Bi-Lateral Migration Database on international long-term migration and from the United Nations High Commissioner for Refugees on the movement of refugees between countries (red circles) were used as sources of international mobility within Africa. Census data (including population-level census, census microdata, and health and demographic surveillance surveys) from 14 countries (green circles) were used as empirical sources of subnational mobility at varying spatial resolutions across countries. Call details records (CDR) were used to quantify subnational mobility patterns at a fine temporal and spatial resolution and were available for five countries (purple circles). Tailored interviews at specific sites (black circles) were also used to quantify mobility. Social media usage at a fine spatial resolution (yellow circles) from two time periods were used to characterise mobility patterns in one country. (b) Locations for which human mobility proxies derived from empirical data shown in (a) have been used in the literature. Subnational mobility proxies for 44 countries in Africa (blue circles) were generated using census, CDR, or both, using data from one or more of 14 African countries where these data were available. For 37 countries in Africa, mobility proxies were the only source of information on subnational mobility. (c) Locations for which mobility proxies were derived either from data sources not shown in (a) or indirectly from other mobility proxies. The former were obtained through mobility models calibrated to empirical data from countries outside Africa (orange circles, flight data from Europe, the United States, and Canada), or from indirect evidence of mobility such as disease spread (pink circle). The latter were characterised using mobility proxies from a different country (cyan circles). In all panels, the size of the open circles indicates the spatial scale of the data or proxy, with smaller circles indicating higher spatial resolution. The number of empirical data sets or mobility proxies derived from a given source is indicated in parenthesis in the legends.

Out of 54 countries, we found only 17 with subnational mobility data. These were informed by census (n = 14 countries), mobile phone records (CDR, n = 5, Côte d’Ivoire, Kenya, Namibia, Sierra Leone, and Senegal), social media records (n = 1) [49] or dedicated surveys (n = 5) [50, 51]. Censuses include surveys designed to measure changes in socio-demographic trends (such as internal migration) in a country, e.g. recording a change in address at ADM1 level over the last 1, 5, or 15 year period. These data are made available as either aggregate statistics (referred to as census or population-level census) or individual records (referred to as census microdata or individual-level census). Harmonised census microdata across different countries provided by the Integrated Public Use Microdata Series [52] were frequently used to quantify subnational mobility. Individual-level census data from specific geographic locations (rather than the entire country) were also available and informed more temporally and spatially resolved movements [53–55]. CDR provided the most spatially and temporally resolved data and were used for methodological research [56–58] as well as research into infectious diseases including Ebola [59], malaria [60, 61], schistosomiasis [62], and cholera [63]. Similarly, mobility data from social media use (i.e. geolocated tweets) were derived at a high spatial resolution (ADM3 in South Africa) [49]. Dedicated surveys collected mobility data relevant to the spread of a specific disease in a specific location [50, 51].

### Mobility proxies

The empirical data described above were used to derive subnational mobility proxies for 44 of 54 countries in Africa, 37 of which had no empirical data informing subnational mobility (Fig. 2b). These proxies were often generated using mobility models fitted to census microdata from one or more countries, or CDR data from other African countries [4, 64, 65]. We also identified mobility proxies estimated from flight data from Europe and North America, and subsequently used to inform mobility between 140 cities across 43 countries in Africa (Fig. 2c) [66]. Overall, we identified subnational mobility proxies for 51 of 54 countries.

We found that typically, mobility proxies were not only generated using data from different locations but also extrapolated the mobility patterns from empirical data to earlier or later time periods, sometimes over a decade apart [4, 61, 63, 65, 67].

In addition to those relying on empirical movement data, mobility proxies were also inferred from indirect evidence such as spatio-temporal trends in disease incidence [68–70] or pathogen genomic information [71, 72] (Fig. 2c).

Finally, mobility proxies were sometimes used to model human movement in different countries or at a different spatial scale, e.g. using gravity model parameters fitted to ADM1 unit data to describe movements between ADM2 units, in the same or another country [73].

Typically, mobility proxies quantified movement information as absolute flows over a specified time window (i.e. the number of people moving between a source and destination) [4, 64–66, 68, 69, 72], or relative flows (i.e. probability of moving from a source to a given destination, conditional on moving out of the source) [51, 57, 60, 63]. Some studies focusing on disease spread also characterised directly the probability of transmission of a pathogen between locations in a given time unit [71, 73]. The focus was therefore to quantify the movement between locations and not the probability of remaining in the same place. However, this assumption was rarely stated explicitly, e.g. clarifying that relative flows are conditional on moving in the first place. In fact, we identified only one study that estimated the probability of not moving out of a spatial unit over an year (i.e. *p*^*stay*^), albeit at a gross temporal scale [62]

In studies that used empirical data sources to calibrate mobility models, the underlying data were rarely shared with the publication, even when commercial restrictions did not prevent data sharing (55/150 empirical data sets readily available). Mobility proxies were more often available (125/168) [64–66, 73].

### Epidemic Simulation Study

We illustrate the potential implications arising from the use of mobility proxies in epidemic models. We present results for our main scenario in which mobility proxies for France are generated from a gravity model calibrated to Portuguese movement data with 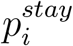 set as the average across all Portuguese administrative units (and vice versa for Portugal). We compare each summary metric (see Methods) for simulations using mobility proxies against a baseline (using observed mobility data).

### Performance of mobility models

The mobility proxies fitted the observed movement data moderately well in both Portugal and France (*R*^2^ = 0.44 and 0.47 respectively, Fig. 3A, Fig. 4A). In a scenario where the mobility proxy was based on data from the same country, the fit was marginally better (*R*^2^ = 0.48 and 0.55 respectively), and only slightly decreased when using 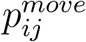 fitted to the other country and a local 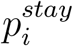 (Suppl Tab. S4). These results suggest that, in both countries, the mobility model we used (see Eqs. (2) to (4)) is only able to explain about half of the variance in the mobility data, and performance decreases by about 10% when fitted to data from another country.

**Figure 3:**
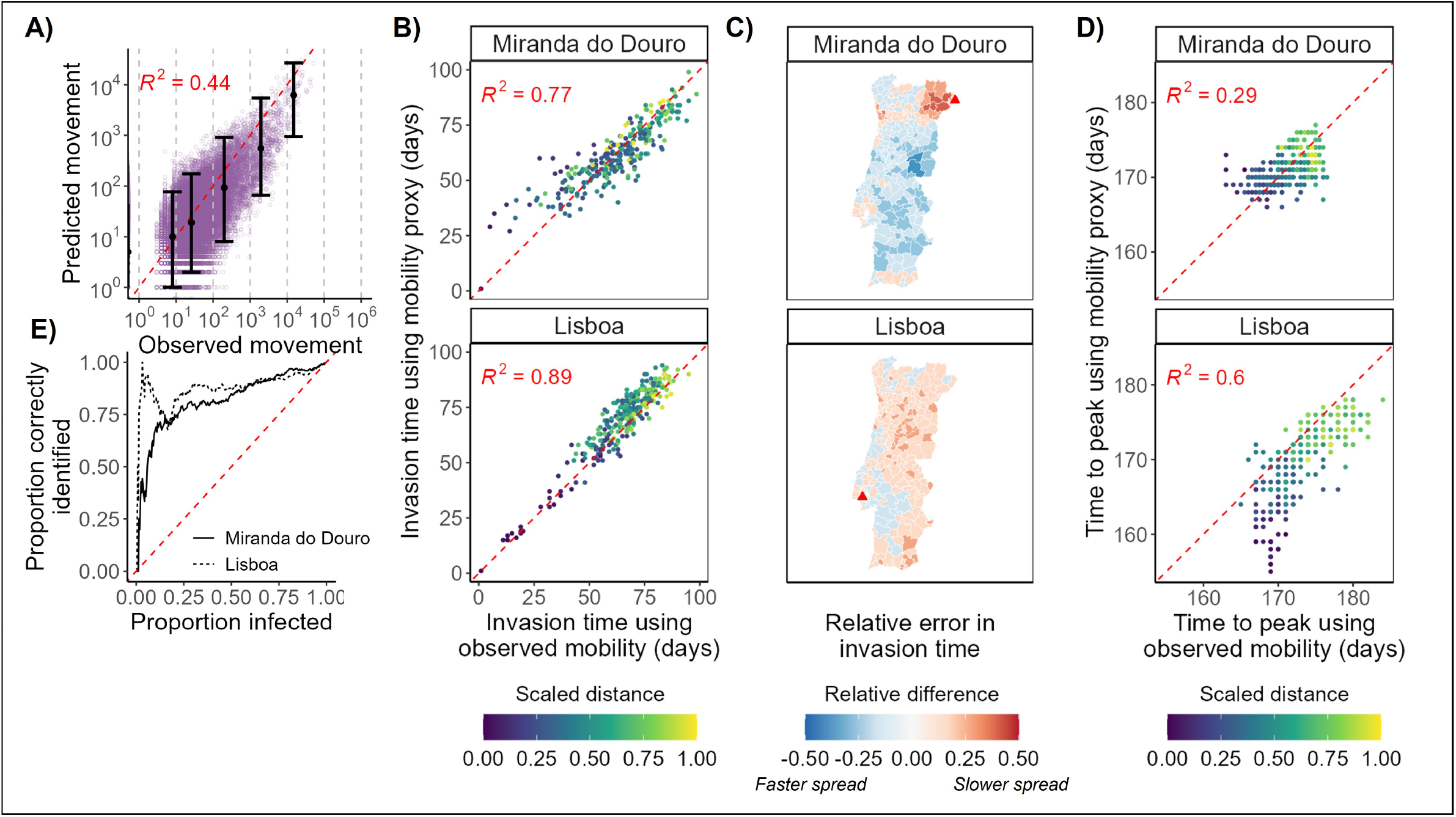
Mobility predictions and epidemic simulations for Portugal. **A)** Predicted numbers moving between Portuguese ADM2 units vs observed numbers from mobile phone call records. Each purple dot represents a pair of ADM2 units. The black dots show the median predicted value (y value) versus the median observed value in each bin (where the bins are marked with grey dashed lines). Error bars show the 2.5% and 97.5% quantiles of the predicted values in that bin. **B) and D)** show respectively the median invasion time 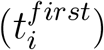 and the median peak time 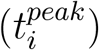 amongst residents of each spatial unit (represented by a dot) across all simulations where an epidemic was successfully seeded. Plots compare the times when using observed mobility patterns (x-axis) vs the mobility proxy (y-axis) from our central scenario in the epidemic model (see Methods). The colour represents the normalised distance of each patch from the seed location (shown by the title of each grid), calculated by dividing the distance from the seed by the maximum distance from that seed. The red dashed line is where y = x. **C)** Maps of the relative error in invasion time in each spatial unit when using the mobility proxy vs observed data. Blue shading indicates the invasion time occurred earlier when using the mobility proxy, while red shading indicates a later invasion time when using mobility proxies. We use a relative scale, calculated as the difference between invasion time using mobility proxy and in the baseline (using the empirical mobility data), divided by the largest invasion times across all locations when using the proxy. The seeding location is marked with a red triangle. **E)** Invasion order similarity 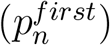. The y-axis shows the proportion of the first *n* patches invaded in an epidemic model using observed mobility that were also among the first *n* locations invaded when a mobility proxy was used in the epidemic model. *n* is the proportion shown on the x-axis. The two lines show different seed locations. The dashed red line shows the expected value if locations are chosen randomly.

**Figure 4:**
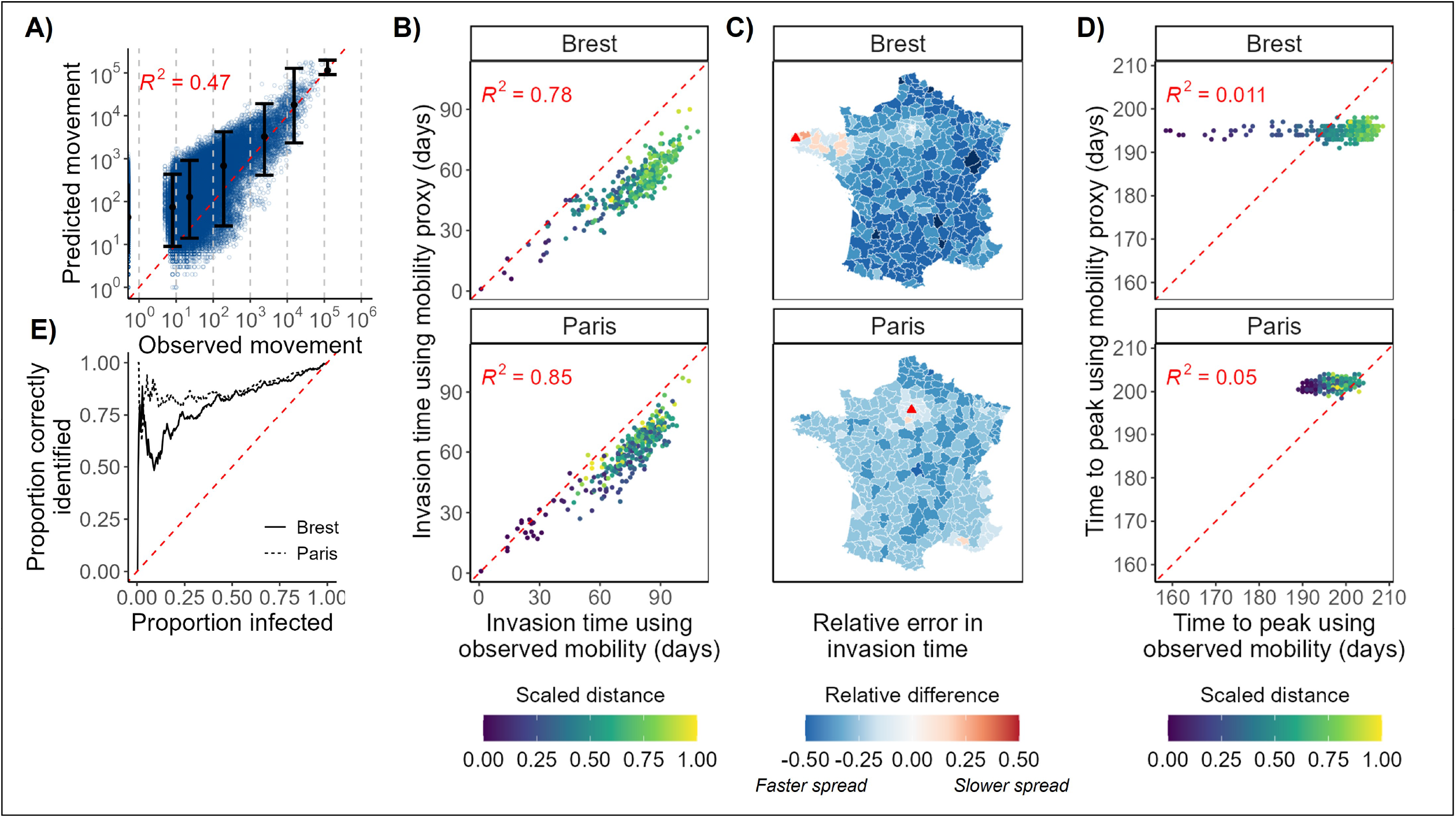
Mobility predictions and epidemic simulations for France. Figure details as in Fig. 4 except here **A)** shows predicted numbers moving between French ADM3 units vs observed numbers from mobile phone call records. Each blue dot represents a pair of ADM3 units.

### Impact on epidemic dynamics

Invasion times 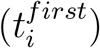 in simulations using mobility proxies were strongly correlated with those using observed movement, especially for epidemics seeded in well-connected locations (*R*^2^ = 0.89 and 0.85 for Lisboa and Paris respectively, and 0.77 and 0.78 for Miranda do Douro and Brest respectively, Fig. 3B, Fig. 4B). However invasion times tended to be under-estimated when using mobility proxies, particularly in France and more so in epidemics with peripheral seeding (Fig. 3C, Fig. 4C). In the most extreme scenario (epidemics seeded in Brest), the invasion times were predicted approximately a month too early (Fig. 4B), a considerable mismatch given the short assumed generation time (the time between infection of a case and their infector) of on average 3 days.

Despite this overall trend to under-predict invasion times, locally around peripheral seeding locations, invasion times were paradoxically over-predicted when using mobility proxies. This led to poor characterisation of the early invasion dynamics, particularly with peripheral seeding (Fig. 3E, Fig. 4E). For epidemics seeded in Brest and Miranda do Douro, only 60% and 55% of the first 20 patches invaded were correctly identified when using mobility proxies, but this increased to 85% and 90% for epidemics seeded in Paris and Lisboa.

The ability to predict epidemic peak times in each spatial unit 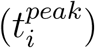 when using mobility proxies was very poor in France (*R*^2^ = 0.05 and 0.01 for Paris and Brest respectively, Fig. 4D) but better in Portugal (*R*^2^ = 0.29 and 0.60 for Miranda do Douro and Lisboa respectively, Fig. 3D). The baseline epidemics seeded in Portugal and in Paris had limited variability in the peak times across administrative units (2-3 weeks). Using mobility proxies reproduced this small amount of variation for Portugal, but not for Paris where a synchronous peak was predicted (1 week). Similarly, although the baseline epidemic seeded in Brest had substantial variability in epidemic peaks (up to 1.5 months apart), again a synchronous peak (1 week) was predicted when using mobility proxies. The sizes of the epidemic peaks were similar across the different movement scenarios.

In both countries, and regardless of the data used to fit the mobility model, the mobility proxies had similar performance in predicting movement data. However, the resulting impact on epidemic dynamics was very different between the two countries. In Portugal, again, epidemic metrics did not differ substantially depending on the data used to fit the mobility model (Suppl Tab. S4, Figs. S5 to S7). In France, using mobility proxies calibrated to Portugal data led to underestimated invasion times, poor invasion order prediction (especially in Brest), and false synchrony in the peaks. However, when using 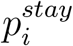 from France, all metrics substantially improved, irrespective of the country used to inform 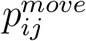 (Suppl Tab. S4, Figs. S1 to S3).

Using the radiation model instead of the gravity model in the main scenario had a mixed impact on epidemic dynamics (Figs. S4 and S8). Overall, the performance was slightly better in France and worse in Portugal. The radiation model improved some metrics, such as the ability to predict early peak timings of the epidemics seeded in Brest, although the later peaks were predicted less well. Use of the radiation model introduced substantial heterogeneity to the Portuguese simulations that was not seen in either the baseline scenario or the additional scenarios using gravity models.

Our results demonstrate that using mobility proxies can have a substantial impact on the predicted epidemic dynamics, with complex and non-intuitive biases, which cannot be predicted when simply comparing the mobility proxies to observed movement data.

## Discussion

Accounting for over 14% of the world’s population [74] and approximately 2.4% of the global airline passenger volume [75], Africa is a large source and sink of national and international movement of populations, and is estimated to bear half of the global burden of infectious diseases [76]. Our review revealed major gaps in the availability of empirical measurements of human mobility in Africa. This leads to mobility proxies being used in place of data, which can impair our understanding of infectious disease dynamics in Africa, as shown by our simulation study.

Although we identified empirical mobility data in all 54 African countries (Fig. 2), data informing subnational movement was only available for 17 countries. These were mostly census data and focused on long term movement (1-5 years), which would only be relevant to the spread of pathogens with very slow progression (e.g. HIV). Despite evidence that these long term movements correlate with short-term mobility [58], census data are either not available at all or infrequently updated in many African countries (Fig. 2).

Highly temporally and spatially resolved mobility data sources [77–79], including but not limited to mobile phone records (CDR data), are increasingly used to characterise human movement and epidemic spread, most frequently in high income countries [21, 43, 80–83]. However, such data is sparse for Africa: we only found 5 African countries with available CDR data, which was subsequently used to derive mobility proxies and examine epidemic spread in 17 other African countries, and which were potentially vastly different in size, population, topography and other factors expected to influence movement patterns.

Given the lack of appropriately resolved mobility data, such use of mobility proxies, typically based on data from other countries, is very common in the African context: we identified subnational mobility proxies for 53/54 African countries. Those were sometimes based on flight data from Europe and America, highlighting the scarcity of reliable local movement data. It is worth noting that mobility proxies often focus on describing absolute or relative flows between distinct locations, but rarely attempt to quantify 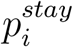, i.e. the number or proportion of people who do not move. This is a critical ingredient to model epidemic spread, where one needs to characterise movement but also the lack thereof.

Our work shows that mobility proxies are imperfect descriptions of empirical mobility, explaining only about half of the variability in the two mobility datasets we considered. Our simulation study demonstrates that this can have substantial and non-intuitive implications on our ability to predict epidemic spread. We focused on the early invasion dynamics and the local peak dynamics, as these metrics would be critical for informing policy, through timely, appropriately scaled and optimally targeted allocation of resources and implementation of control measures [84–86].

In most scenarios explored, predicted invasion times using mobility proxies tended to be earlier than when using empirical mobility data. While this may seem less problematic than the reverse, it could lead to dismissing preventive interventions, wrongly perceived to take too long to implement given the predicted speed of invasion (e.g. building a new healthcare facility). Although the early invasion order was relatively well characterised for epidemics seeded in the capital cities, this was not the case when the epidemics were seeded in peripheral locations, and could lead to inappropriate spatial targeting of control measures. Heterogeneity in local epidemic peak times was overall well quantified in Portugal but not at all for France. Better characterising the peak heterogeneity may help better coordination of resources among different regions, e.g. allowing movement of medical staff, patients or material depending on the predicted local times of maximal epidemic burden.

These issues were overall more apparent in France then Portugal, and were much improved when using 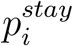 from the correct country. Although 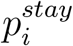 has received little attention by researchers, our work emphasises the importance of collecting data to estimate this parameter in all countries and at a fine spatial resolution, which can be later aggregated if needed. Indeed 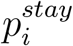 will evidently depend on the size of the spatial unit under consideration. For instance, ADM2 units in Portugal are on average smaller than ADM3 units in France, leading to the aforementioned underestimation of invasion times. Such data collection effort, to characterise nonmovement, may be even more critical than collecting data on the destinations of the movements; indeed results from our epidemic simulations were much more sensitive to changes in 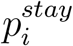 than in 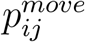.

In our study, we found that using French data to inform Portugal mobility fared much better than the opposite. Population mobility in a country potentially depends on a large number of factors such as the geography of the country, population density, demographics, and distribution of economic opportunities [87–89]. Understanding the key determinants may allow us to predict to what extent and in which contexts 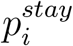 and 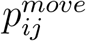 may be informed by data from other countries. However, this would only be possible if highly resolved mobility data from many countries were available, which would reduce the need for mobility proxies. Although in our simulation study, we generated mobility proxies using data from one other country, we note that some popular published proxies [64, 65] use data from multiple countries. It is unclear what the implications for disease modelling are, and whether such data pooling across countries is more appropriate than using data from a single country.

Our work has some limitations. First, since our search strategy was limited to identifying studies that used movement data in mobility models, we may have overlooked studies that primarily described mobility data and did not use any models. However, we believe it is unlikely that we have missed important data sets, as those would have been used by modelling studies captured in our search.

Second, we carried out the search in 2018. Since then, novel sources of mobility data have emerged specifically in the context of the COVID-19 pandemic. This includes community mobility reports from Google and Apple [90, 91]; however those only measure overall mobility trends which cannot directly be used to inform O-D matrices. Furthermore, many such data are being released specifically to support the COVID-19 response, and cannot replace much needed highly-resolved representative and regularly updated mobility data.

Third, we used gravity and radiation mobility models, and a multipatch compartmental model stratified by home and work location, guided by the available mobility data. More sophisticated models of mobility [92] or disease propagation [46, 93–95] could display more complex dynamics. However our simple and parsimonious approach was sufficient to highlight potential biases in epidemic spread predictions stemming from using mobility proxies instead of empirical data; this would only be magnified by using more sophisticated models.

Finally, one aspect we did not consider, which is rarely acknowledged, is that mobility proxies are outputs of statistical models, and hence carry inherent uncertainty. Such uncertainty is rarely quantified, or reported, and almost never propagated in subsequent analysis, e.g. epidemic models.

Overall, our work underscores the need for regularly updated empirical measurements of population movement within and between countries. Despite potential biases in mobile phone usage due to limited and unevenly distributed smartphone ownership [96], such data sources remain the most promising to characterise shortterm human movement at high resolution. Mobile phone operators could consider periodic release of aggregate data sets to support public health efforts. Even though we did not restrict our search to studies related to infectious diseases, most of the included publications focused on their spread, highlighting the centrality of human movement in infectious disease epidemiology. A few data sources underpinned wideranging research on multiple diseases such as cholera, ebola, malaria, and schistosomiasis. This suggests that availability of empirical mobility data could pay large dividends in pandemic preparedness as well as an improved understanding of spread of diseases that are endemic or lead to recurring epidemics [97].

## Supporting information

Supplementary Information 2 - data from scoping review

## Data Availability

Data from the study are available online at https://github.com/j-wardle/mobility_africa_models

## Acknowledgements

We thank Michele Tizzoni and colleagues for publicly sharing data on commuter patterns in France and Portugal, as well as for providing us with additional data on the numbers of people that do not commute. This study is partially funded by the National Institute for Health Research (NIHR) Health Protection Research Unit in Modelling and Health Economics, a partnership between UK Health Security Agency (UKHSA) and Imperial College London, in collaboration with LSHTM (grant code NIHR200908); and acknowledges funding from the MRC Centre for Global Infectious Disease Analysis (reference MR/R015600/1), which is jointly funded by the UK Medical Research Council (MRC) and the UK Foreign, Commonwealth & Development Office (FCDO), under the MRC/FCDO Concordat agreement and is also part of the EDCTP2 programme supported by the European Union; and acknowledges funding by Community Jameel. JW acknowledges research funding from the Wellcome Trust (grant 102169/Z/13/Z). SB acknowledges funding from the Wellcome Trust (grant 219415). Disclaimer: The views expressed are those of the author(s) and not necessarily those of the NIHR, UKHSA or the Department of Health and Social Care. The funders had no role in study design, data collection and analysis, decision to publish, or preparation of the manuscript. For the purpose of open access, the author has applied a ‘Creative Commons Attribution’ (CC BY) licence to any Author Accepted Manuscript version arising from this submission.

## Supplementary information

We share two supplementary files:

1. Supplementary information on the methods and results of the simulation study
2. Data extracted from studies included in the scoping review

## References

[1] S. Merler et al. “Spatiotemporal spread of the 2014 outbreak of Ebola virus disease in Liberia and the effectiveness of non-pharmaceutical interventions: a computational modelling analysis”. In: The Lancet Infectious Diseases 15.2 (2015), pp. 204–211.

[2] W. Yang et al. “Transmission network of the 2014–2015 Ebola epidemic in Sierra Leone”. In: Journal of The Royal Society Interface 12.112 (2015), p. 20150536.

[3] I. Dorigatti et al. “International risk of yellow fever spread from the ongoing outbreak in Brazil, December 2016 to May 2017”. In: Eurosurveillance 22.28 (2017), p. 30572.

[4] M. U. Kraemer et al. “Spread of yellow fever virus outbreak in Angola and the Democratic Republic of the Congo 2015–16: a modelling study”. In: The Lancet infectious diseases 17.3 (2017), pp. 330–338.

[5] A. J. Tatem et al. “Global transport networks and infectious disease spread”. In: Advances in parasitology 62 (2006), pp. 293–343. issn: 2163-60790065-308X. doi: 10.1016/S0065-308X(05)62009-X. url: https://pubmed.ncbi.nlm.nih.gov/16647974%20https://www.ncbi.nlm.nih.gov/pmc/articles/PMC3145127/.

[6] A. Findlater and I. I. Bogoch. “Human Mobility and the Global Spread of Infectious Diseases: A Focus on Air Travel”. In: Trends in parasitology 34.9 (2018), pp. 772–783. issn: 1471-5007 1471-4922. doi: 10.1016/j.pt.2018.07.004. url: https://pubmed.ncbi.nlm.nih.gov/30049602%20https://www.ncbi.nlm.nih.gov/pmc/articles/PMC7106444/.

[7] J. Li et al. “The emergence, genomic diversity and global spread of SARS-CoV-2”. In: Nature (2021), pp. 1–11.

[8] Á. O’Toole et al. “Tracking the international spread of SARS-CoV-2 lineages B. 1.1. 7 and B. 1.351/501Y-V2”. In: Wellcome Open Research 6 (2021).

[9] R. Viana et al. “Rapid epidemic expansion of the SARS-CoV-2 Omicron variant in southern Africa”. In: medRxiv (2021).

[10] I. I. Bogoch et al. “Assessment of the potential for international dissemination of Ebola virus via commercial air travel during the 2014 west African outbreak”. In: The Lancet 385.9962 (2015), pp. 29–35.

[11] M. Gilbert et al. “Preparedness and vulnerability of African countries against importations of COVID-19: a modelling study”. In: The Lancet 395.10227 (2020), pp. 871–877.

[12] A. Craig et al. “Risk of COVID-19 importation to the Pacific islands through global air travel”. In: Epidemiology & Infection 148 (2020).

[13] M. Chinazzi et al. “The effect of travel restrictions on the spread of the 2019 novel coronavirus (COVID-19) outbreak”. In: Science 368.6489 (2020), p. 395. doi: 10.1126/science.aba9757. url: http://science.sciencemag.org/content/368/6489/395.abstract.

[14] T. D. Hollingsworth et al. “Will travel restrictions control the international spread of pandemic influenza?” In: Nature medicine 12.5 (2006), pp. 497–499.

[15] A. R. Tuite et al. “Cholera epidemic in Haiti, 2010: using a transmission model to explain spatial spread of disease and identify optimal control interventions”. In: Annals of internal medicine 154.9 (2011), pp. 593–601.

[16] J. T. Wu et al. “Spatial considerations for the allocation of pre-pandemic influenza vaccination in the United States”. In: Proceedings of the Royal Society B: Biological Sciences 274.1627 (2007), pp. 2811–2817.

[17] I. M. Longini et al. “Containing pandemic influenza at the source”. In: Science 309.5737 (2005), pp. 1083–1087.

[18] A. J. Kucharski et al. “Measuring the impact of Ebola control measures in Sierra Leone”. In: Proceedings of the National Academy of Sciences 112.46 (2015), pp. 14366–14371.

[19] A. J. Tatem. “Mapping population and pathogen movements”. In: International Health 6.1 (2014), pp. 5–11. issn: 1876-3413. doi: 10.1093/inthealth/ihu006. url: https://doi.org/10.1093/inthealth/ihu006.

[20] M. U. G. Kraemer et al. “Progress and Challenges in Infectious Disease Cartography”. In: Trends in Parasitology 32.1 (2016), pp. 19–29. issn: 1471-4922. doi: https://doi.org/10.1016/j.pt.2015.09.006. url: http://www.sciencedirect.com/science/article/pii/S147149221500207X.

[21] B. Jeffrey et al. “Anonymised and Aggregated Crowd Level Mobility Data from Mobile Phones Suggests That Initial Compliance with COVID-19 Social Distancing Interventions Was High and Geographically Consistent across the UK”. In: Wellcome Open Research 5 (July 2020), pp. 170–170. doi: 10.12688/wellcomeopenres.15997.1.

[22] N. W. Ruktanonchai et al. “Assessing the impact of coordinated COVID-19 exit strategies across Europe”. In: Science 369.6510 (2020), pp. 1465–1470.

[23] A. Ascani et al. “Mobility in Times of Pandemics: Evidence on the Spread of COVID19 in Italy’s Labour Market Areas”. In: Structural Change and Economic Dynamics 58 (Sept. 2021), pp. 444–454. issn: 0954349X. doi: 10.1016/j.strueco.2021.06.016.

[24] T. Ramiadantsoa et al. “Existing human mobility data sources poorly predicted the spatial spread of SARS-CoV-2 in Madagascar”. In: Epidemics 38 (2022), p. 100534.

[25] A. Wesolowski et al. “Connecting mobility to infectious diseases: the promise and limits of mobile phone data”. In: The Journal of infectious diseases 214.uppl 4 (2016), S414–S420.

[26] A. Wesolowski et al. “Heterogeneous mobile phone ownership and usage patterns in Kenya”. In: PloS one 7.4 (2012), e35319.

[27] A. Wesolowski et al. “The impact of biases in mobile phone ownership on estimates of human mobility”. In: Journal of the Royal Society Interface 10.81 (2013), p. 20120986.

[28] S. Lai et al. “Measuring mobility, disease connectivity and individual risk: a review of using mobile phone data and mHealth for travel medicine”. In: Journal of travel medicine 26.3 (2019), taz019.

[29] M. U. G. Kraemer et al. “Utilizing General Human Movement Models to Predict the Spread of Emerging Infectious Diseases in Resource Poor Settings”. In: Scientific Reports 9.1 (Dec. 2019), p. 5151. issn: 2045-2322. doi: 10.1038/s41598-019-41192-3.

[30] G. K. Zipf. “The P 1 P 2/D hypothesis: on the intercity movement of persons”. In: American sociological review 11.6 (1946), pp. 677–686.

[31] F. Simini et al. “A universal model for mobility and migration patterns”. In: Nature 484.7392 (2012), pp. 96–100. issn: 1476-4687. doi: 10.1038/nature10856. url: https://doi.org/10.1038/nature10856.

[32] S. Bhatia et al. “Using Digital Surveillance Tools for near Real-Time Mapping of the Risk of Infectious Disease Spread”. In: NPJ Digital Medicine 4.1 (Apr. 2021), pp. 1–10. doi: 10.1038/s41746-021-00442-3.

[33] History of Ebola Virus Disease (EVD) Outbreaks. url: https://www.cdc.gov/vhf/ebola/history/chronology.html (visited on 02/28/2022).

[34] Yellow Fever: Recent Outbreaks in Africa. url: https://africacdc.org/disease/yellow-fever/ (visited on 02/28/2022).

[35] Cholera: Recent Outbreaks in Africa. url: https://africacdc.org/disease/cholera/ (visited on 02/28/2022).

[36] Measles: Recent Outbreaks in Africa. url: https://africacdc.org/disease/measles/ (visited on 02/28/2022).

[37] A. C. Tricco et al. “PRISMA Extension for Scoping Reviews (PRISMA-ScR): Checklist and Explanation”. In: Annals of Internal Medicine 169.7 (Oct. 2018), pp. 467–473. issn: 0003-4819, 1539-3704. doi: 10.7326/M18-0850.

[38] C. Viboud et al. “Synchrony, waves, and spatial hierarchies in the spread of influenza”. In: science 312.5772 (2006), pp. 447–451.

[39] R. M. Eggo et al. “Spatial dynamics of the 1918 influenza pandemic in England, Wales and the United States”. In: Journal of the Royal Society Interface 8.55 (2011), pp. 233–243.

[40] V. Charu et al. “Human mobility and the spatial transmission of influenza in the United States”. In: PLoS computational biology 13.2 (2017), e1005382.

[41] J. Truscott and N. M. Ferguson. “Evaluating the adequacy of gravity models as a description of human mobility for epidemic modelling”. In: (2012).

[42] N. Bharti et al. “Measles on the edge: coastal heterogeneities and infection dynamics”. In: PloS one 3.4 (2008), e1941.

[43] M. Tizzoni et al. “On the Use of Human Mobility Proxies for Modeling Epidemics”. In: PLOS Computational Biology 10.7 (2014), e1003716. doi: 10.1371/journal.pcbi.1003716. url: https://doi.org/10.1371/journal.pcbi.1003716

[44] C. Song et al. “Limits of predictability in human mobility”. In: Science 327.5968 (2010), pp. 1018–1021.

[45] S. Riley et al. “Five challenges for spatial epidemic models”. In: Epidemics 10 (2015), pp. 68–71.

[46] M. J. Keeling et al. “Individual identity and movement networks for disease metapopulations”. In: Proceedings of the National Academy of Sciences 107.19 (2010), pp. 8866–8870.

[47] Ç. Özden et al. “Where on Earth Is Everybody? The Evolution of Global Bilateral Migration 1960–2000”. In: The World Bank Economic Review 25.1 (Jan. 2011), pp. 12–56. issn: 1564-698X, 0258-6770. doi: 10.1093/wber/lhr024.

[48] The UN Refugee Agency Refugee Data Finder. url: https://www.unhcr.org/refugee-statistics/ (visited on 02/24/2022).

[49] A. Dobra, R. Mohammadi, et al. “Loglinear model selection and human mobility”. In: The Annals of Applied Statistics 12.2 (2018), pp. 815–845.

[50] J. O. Yukich et al. “Travel history and malaria infection risk in a low-transmission setting in Ethiopia: a case control study”. In: Malaria Journal 12.1 (2013), p. 33.

[51] J. M. Marshall et al. “Mathematical models of human mobility of relevance to malaria transmission in Africa”. In: Scientific Reports 8.1 (2018), p. 7713.

[52] Integrated Public Use Microdata Series, International: Version 7.3[dataset. url: https://international.ipums.org/international/index.shtml (visited on 02/24/2022).

[53] M. A. Collinson et al. “Migration and the epidemiological transition: insights from the Agincourt sub-district of northeast South Africa”. In: Global Health Action 7.1 (2014), p. 23514.

[54] A. Dobra et al. “Space-time migration patterns and risk of HIV acquisition in rural South Africa”. In: AIDS (London, England) 31.1 (2017), p. 137.

[55] J. R. Andrews et al. “Projecting the Benefits of Antiretroviral Therapy for HIV Prevention: The Impact of Population Mobility and Linkage to Care”. In: The Journal of Infectious Diseases 206.4 (2012), pp. 543–551.

[56] X. Lu et al. “Approaching the Limit of Predictability in Human Mobility”. In: Scientific Reports 3 (2013), p. 2923.

[57] J. T. Matamalas et al. “Assessing reliable human mobility patterns from higher order memory in mobile communications”. In: Journal of The Royal Society Interface 13.121 (2016), p. 20160203.

[58] A. Wesolowski et al. “The use of census migration data to approximate human movement patterns across temporal scales”. In: PloS One 8.1 (2013), e52971.

[59] C. M. Peak et al. “Population mobility reductions associated with travel restrictions during the Ebola epidemic in Sierra Leone: use of mobile phone data”. In: International journal of epidemiology 47.5 (2018), pp. 1562–1570.

[60] A. M. Tompkins and N. McCreesh. “Migration statistics relevant for malaria transmission in Senegal derived from mobile phone data and used in an agent-based migration model.” In: Geospatial Health 11.1 Supp (2016), p. 408.

[61] N. W. Ruktanonchai et al. “Identifying Malaria Transmission Foci for Elimination Using Human Mobility Data”. In: PLoS Computational Biology 12.4 (2016), e1004846.

[62] L. Mari et al. “Big-data-driven modeling unveils country-wide drivers of endemic schistosomiasis”. In: Scientific reports 7.1 (2017), p. 489.

[63] F. Finger et al. “Mobile phone data highlights the role of mass gatherings in the spreading of cholera outbreaks”. In: Proceedings of the National Academy of Sciences 113.23 (2016), pp. 6421–6426.

[64] A. Sorichetta et al. “Mapping internal connectivity through human migration in malaria endemic countries”. In: Scientific Data 3 (2016), p. 160066.

[65] A. Wesolowski et al. “Commentary: containing the Ebola outbreak-the potential and challenge of mobile network data”. In: PLoS currents 6 (2014).

[66] Z. Huang et al. “An open-access modeled passenger flow matrix for the global air network in 2010”. In: PloS one 8.5 (2013), e64317.

[67] D. K. Pindolia et al. “The demographics of human and malaria movement and migration patterns in East Africa”. In: Malaria Journal 12.1 (2013), p. 397.

[68] J.P. D’Silva and M. C. Eisenberg. “Modeling spatial invasion of Ebola in West Africa”. In: Journal of Theoretical Biology 428 (2017), pp. 65–75.

[69] S. P. Silal et al. “Hitting a moving target: a model for malaria elimination in the presence of population movement”. In: PLoS One 10.12 (2015), e0144990.

[70] A. J. Tatem et al. “Spatial accessibility and the spread of HIV-1 subtypes and recombinants”. In: Aids 26.18 (2012), pp. 2351–2360.

[71] K. B. Gustafson and J. L. Proctor. “Identifying spatio-temporal dynamics of Ebola in Sierra Leone using virus genomes”. In: Journal of The Royal Society Interface 14.136 (2017), p. 20170583.

[72] G. Dudas et al. “Virus genomes reveal factors that spread and sustained the Ebola epidemic”. In: Nature 544.7650 (2017), p. 309.

[73] A. M. Kramer et al. “Spatial spread of the West Africa Ebola epidemic”. In: Royal Society Open Science 3.8 (2016), p. 160294.

[74] Africa Population. url: https://www.worldometers.info/world-population/africa-population/ (visited on 02/03/2022).

[75] Air Traffic Statistics. url: https://www.aci-africa.aero/data-centre/air-traffic-statistics/ (visited on 02/03/2022).

[76] A. Boutayeb. “The Impact of Infectious Diseases on the Development of Africa”. In: Handbook of Disease Burdens and Quality of Life Measures. Ed. by V. R. Preedy and R. R. Watson. New York, NY: Springer New York, 2010, pp. 1171–1188. isbn: 978-0-387-78664-3 978-0-387-78665-0. doi: 10.1007/978-0-387-78665-0_66.

[77] National Travel Survey: 2020. url: https://www.gov.uk/government/statistics/national-travel-survey-2020/national-travel-survey-2020 (visited on 02/03/2022).

[78] Eurostat: your key to European Statistics. url: https://ec.europa.eu/eurostat/web/transport/data/database (visited on 02/03/2022).

[79] Transport for London: Our Open Data. url: https://tfl.gov.uk/info-for/open-data-users/our-open-data (visited on 02/03/2022).

[80] E. Pepe et al. “COVID-19 outbreak response, a dataset to assess mobility changes in Italy following national lockdown”. In: Scientific data 7.1 (2020), pp. 1–7.

[81] S. Silm et al. “Temporary population mobilities between Estonia and Finland based on mobile phone data and the emergence of a cross-border region”. In: European Planning Studies 29.4 (2021), pp. 699–719.

[82] S. Chang et al. “Mobility network models of COVID-19 explain inequities and inform reopening”. In: Nature 589.7840 (2021), pp. 82–87.

[83] F. Schlosser et al. “COVID-19 lockdown induces disease-mitigating structural changes in mobility networks”. In: Proceedings of the National Academy of Sciences 117.52 (2020), pp. 32883–32890.

[84] SPI-M Modelling Summary. 2013. url: https://assets.publishing.service.gov.uk/government/uploads/system/uploads/attachment_data/file/756738/SPI-M_modelling_summary_final.pdf.

[85] L. Danon et al. “A Spatial Model of COVID-19 Transmission in England and Wales: Early Spread, Peak Timing and the Impact of Seasonality”. In: Philosophical Transactions of the Royal Society B: Biological Sciences 376.1829 (July 2021), p. 20200272. issn: 0962-8436, 1471-2970. doi: 10.1098/rstb.2020.0272.

[86] M. Deschepper et al. “Prediction of Hospital Bed Capacity during the COVID-19 Pandemic”. In: BMC Health Services Research 21.1 (Dec. 2021), p. 468. issn: 1472-6963. doi: 10.1186/s12913-021-06492-3.

[87] C. Bonifazi and F. Heins. “Long-term trends of internal migration in Italy”. In: International Journal of Population Geography 6.2 (2000), pp. 111–131.

[88] F. Castelli. “Drivers of migration: why do people move?” In: Journal of Travel Medicine 25 (1 2018).

[89] A. J. Conlan et al. “Human mobility data from the BBC Pandemic project”. In: medRxiv (2021).

[90] COVID-19 Community Mobility Reports. url: https://www.google.com/covid19/mobility/ (visited on 02/03/2022).

[91] COVID-19 Mobility Trends Reports. url: https://covid19.apple.com/mobility (visited on 02/03/2022).

[92] H. R. Meredith et al. “Characterizing Human Mobility Patterns in Rural Settings of Sub-Saharan Africa”. In: eLife 10 (Sept. 2021). Ed. by J. Flegg et al., e68441. issn: 2050-084X. doi: 10.7554/eLife.68441.

[93] K. Van Kerckhove et al. “The impact of illness on social networks: implications for transmission and control of influenza”. In: American journal of epidemiology 178.11 (2013), pp. 1655–1662.

[94] D. J. Haw et al. “Differential mobility and local variation in infection attack rate”. In: PLoS computational biology 15.1 (2019), e1006600.

[95] K. Prem et al. “Projecting social contact matrices in 152 countries using contact surveys and demographic data”. In: PLoS computational biology 13.9 (2017), e1005697.

[96] The Mobile Economy Sub-Saharan Africa 2021. url: https://www.gsma.com/mobileeconomy/wp-content/uploads/2021/09/GSMA_ME_SSA_2021_English_Web_Singles.pdf (visited on 02/03/2022).

[97] C. O. Buckee et al. “Aggregated mobility data could help fight COVID-19”. In: Science 368.6487 (2020), pp. 145–146.

