## Supplementary Information 1 for "Gaps in mobility data and implications for modelling epidemic spread: a scoping review and simulation study"

#### Contents

**1 Overview** **1**

**2 Methods** **1**

**3 Summary of location and movement information** **3**

**4 Supplementary results** **4**

### 1 Overview

In this document, we present supplementary information on the methodology and results of our simulation study. In Section 2 we describe how mobility proxies were generated for the non-baseline scenarios explored in our analysis, and provide further information on the processing of movement data and the epidemic model. Section 3 provides the key geographic characteristics of the settings for the simulation study, while Section 4 presents the results from epidemic simulations for the range of additional scenarios not shown in the main text.

### 2 Methods

The information in this section provides complementary details to the Methods section of the main text.

#### Data sources

The generation of mobility proxies for France and Portugal required data on population sizes and distances between administrative units (or "locations"). The populations of each location $i$ ($pop_i$) were derived using population data from WorldPop [1, 2]. The distances between locations $i$ and $j$ ($d_{ij}$) were measured between the population-weighted centroids of each administrative unit, computed from WorldPop population data [1, 2] and GADM shapefiles of administrative boundaries [3].

#### Software

All epidemic simulations and analyses were performed using the R software [4]. We fitted gravity and radiation models to observed data using the *movement* package in R [5].

#### Mobility proxies

In addition to the central scenario described in the main text, we generated four other mobility proxies for France and Portugal by using different combinations of data sources to inform $p_i^{stay}$ and $p_{ij}^{move}$ (see Table S1). The data were either from the same country as the setting for which we generated a proxy (referred to as *local*) or from the alternative country (for which we use the notation *alt*). In the case of $p_i^{stay}$, *alt* means that the average value across all locations from the other country was used in all locations. When using the *local* data source, the observed $p_i^{stay}$ for each individual location was used.

In Suppl. Scenario 3 Table S1, we generated a mobility proxy in which *local* data was used but at a more aggregated spatial scale (for which we use the notation *local*∗) than the scale at which we were modelling the disease dynamics. For France, this meant that the $p_i^{stay}$ value used for ADM3 spatial units was based on the average value observed across all of the ADM2 spatial units in France. For $p_{ij}^{move}$ the gravity model was fitted to the observed movement data between ADM2 units, with the estimated parameters then used to predict movements between ADM3 units. The same approach was used for the Portuguese data, although here the aggregated spatial scale was ADM1 and the higher resolution level was ADM2.

To explore the robustness of our results to the choice of mobility model, we also considered a scenario similar to our main one (with $p_i^{stay}$ and $p_{ij}^{move}$ informed by data from the other country, i.e. *alt*) but instead using a radiation model to predict $p_{ij}^{move}$. In the radiation model, the flow of people between locations $i$ and $j$ is proportional to:

$$p_{ij}^{move} \propto \frac{N_i N_j}{(N_i + s_{ij})(N_i + N_j + s_{ij})} \tag{S1}$$

where $N_i$ and $N_j$ are the populations living in the origin and destination locations $i$ and $j$ respectively, and $s_{ij}$ is the total population in the circle of radius $r_{ij}$ and centre $i$ (not including $N_i$ and $N_j$) [6].

| Scenario | Source of $p_i^{stay}$ | Source of $p_{ij}^{move}$ | Mobility model used |
|---|---|---|---|
| Main | alt | alt | gravity |
| Suppl. 1 | local | alt | gravity |
| Suppl. 2 | local | local | gravity |
| Suppl. 3 | local* | local* | gravity |
| Suppl. 4 | alt | alt | radiation |

Table S1: **Summary of the mobility proxy scenarios explored in the simulation study**. *alt* indicates that $p_{stay}$ or $p_{move}$ were based on data from a different country. *local* is used when $p_{stay}$ or $p_{move}$ are based on data from the same country. *local** indicates that we use data from the correct country but at the wrong spatial scale.

##### Epidemic model

We simulated epidemics of a flu-like pathogen for which we assumed a mean latent period ($1/\delta$) of 1.0 days, mean infectious period ($1/\gamma$) of 2.0 days and per capita transmission rate ($\beta$) of 0.6 days$^{-1}$ (corresponding to a basic reproduction number of 1.2).

##### Code and data availability

All code used in this analysis is available at `https://github.com/j-wardle/mobility_africa_models`. The simulation study is based on data made available by Tizzoni et al. [7].

#### 3 Summary of location and movement information

Table S2 provides a summary of the population data for locations in France and Portugal, and the average value of $p_i^{stay}$ across all spatial units in the observed data for each country. Table S3 shows the estimated gravity model parameters from fitting to the observed and aggregated data.

| | France | Portugal |
|---|---|---|
| Spatial resolution of observed data | ADM3 | ADM2 |
| Number of mainland spatial units | 323 | 278 |
| Median patch population (min; max) | 122,488 (8,227; 2,179,720) | 15,700 (1,329; 584,079) |
| Mean $p_i^{stay}$ (min; max) | 0.66 (0.19; 0.87) | 0.29 (0.00; 0.80) |

Table S2: Summary of the location data for France and Portugal

|  | **France** | **Portugal** |
|---|---|---|
| **Highest spatial resolution** | ADM3 | ADM2 |
| $\alpha$ | 0.307 | 0.357 |
| $\beta$ | 0.462 | 0.590 |
| $\gamma$ | 1.604 | 1.701 |
| **Aggregated spatial resolution** | ADM2 | ADM1 |
| $\alpha$ | 0.398 | 0.531 |
| $\beta$ | 0.458 | 0.618 |
| $\gamma$ | 1.353 | 1.073 |

Table S3: Estimates for the gravity model parameters from fitting to the commuting data at different spatial resolutions. The top set of parameters show the fit to data at their highest spatial resolutions (i.e the resolution at which the data was collected). This is ADM3 for France and ADM2 for Portugal. The bottom set of parameters are from the fit to data at aggregated spatial scales (ADM2 for France and ADM3 for Portugal).

### 4 Supplementary results

Here we present results for the scenarios not included in the main text. We compare predicted and observed movement values, and the invasion times ($t_i^{first}$) and peak times ($t_i^{peak}$) for epidemic simulations based on a mobility proxy with those based on observed movement (Table S4). Figure panels displaying the results from each scenario follow Table S4.

| **Scenario** | **$R^2$ between** | **France** | **Portugal** |
|---|---|---|---|
| Main | movement | 0.47 | 0.44 |
| | invasion time | Brest - 0.78, Paris - 0.85 | Miranda - 0.77, Lisboa - 0.89 |
| | time to peak | Brest - 0.01, Paris - 0.05 | Miranda - 0.29, Lisboa - 0.60 |
| Suppl. 1 | movement | 0.54 | 0.49 |
| | invasion time | Brest - 0.81, Paris - 0.36 | Miranda - 0.75, Lisboa - 0.87 |
| | time to peak | Brest - 0.72, Paris - 0.36 | Miranda - 0.16, Lisboa - 0.51 |
| Suppl. 2 | movement | 0.55 | 0.48 |
| | invasion time | Brest - 0.84, Paris - 0.87 | Miranda - 0.78, Lisboa - 0.88 |
| | time to peak | Brest - 0.56, Paris - 0.40 | Miranda - 0.17, Lisboa - 0.52 |
| Suppl. 3 | movement | 0.50 | 0.40 |
| | invasion time | Brest - 0.82, Paris - 0.86 | Miranda - 0.65, Lisboa - 0.81 |
| | time to peak | Brest - 0.68, Paris - 0.43 | Miranda - 0.19, Lisboa - 0.31 |
| Suppl. 4 | movement | 0.60 | 0.51 |
| | invasion time | Brest - 0.76, Paris - 0.82 | Miranda - 0.47, Lisboa - 0.87 |
| | time to peak | Brest - 0.67, Paris - 0.39 | Miranda - 0.44, Lisboa - 0.48 |

Table S4: $R^2$ values are shown for the relationships between predicted and observed movement values between any two locations. We also present $R^2$ values between invasion times and peak times when using mobility proxies in epidemic simulations versus observed mobility data. The city names correspond to the location where the epidemics were seeded. See Table S1 for further information on the mobility proxy used in each scenario.

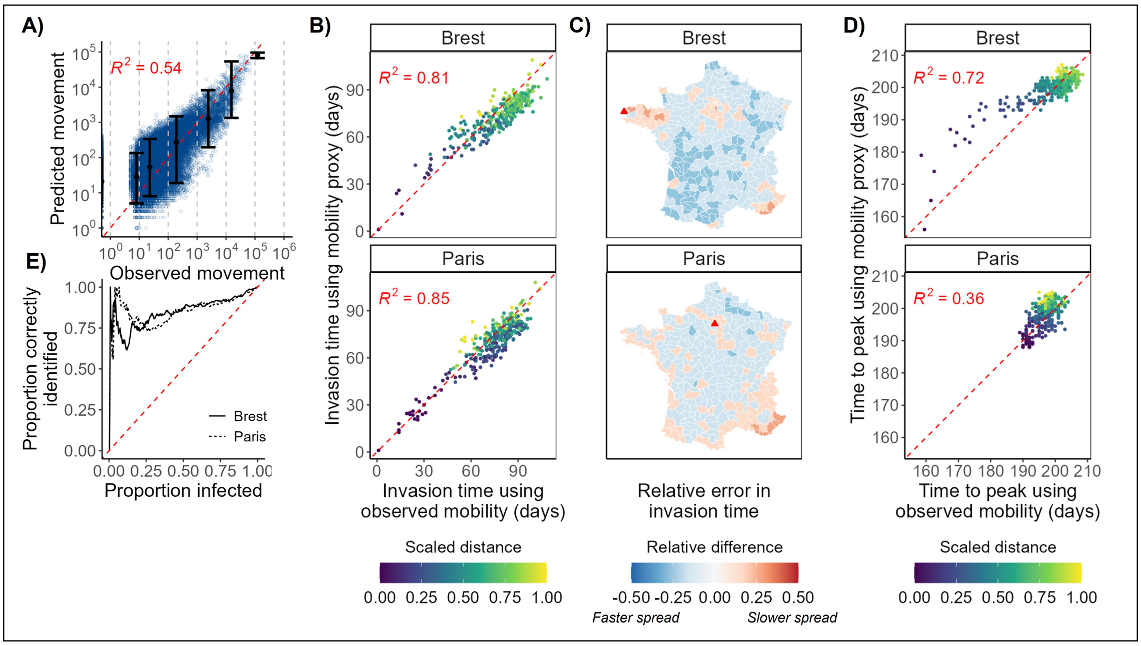


Figure S1: **Mobility predictions and epidemic simulations for France under Suppl. Scenario 1.** In this scenario $p_i^{stay}$ is informed by local data, and $p_{ij}^{move}$ is informed by data from Portugal (see Table S1 for further scenario details). **A)** Predicted numbers moving between French ADM3 units vs observed numbers from mobile phone call records. Each blue dot represents a pair of ADM3 units. The black dots show the median predicted value (y value) versus the median observed value in each bin (where the bins are marked with grey dashed lines). Error bars show the 2.5% and 97.5% quantiles of the predicted values in that bin. **B) and D)** show respectively the median invasion time ($t_i^{first}$) and the median peak time ($t_i^{peak}$) amongst residents of each spatial unit (represented by a dot) across all simulations where an epidemic was successfully seeded. Plots compare the times when using observed mobility patterns (x-axis) vs the mobility proxy (y-axis) from our central scenario in the epidemic model (see Methods). The colour represents the normalised distance of each patch from the seed location (shown by the title of each grid), calculated by dividing the distance from the seed by the maximum distance from that seed. The red dashed line is where y = x. **C)** Maps of the relative error in invasion time in each spatial unit when using the mobility proxy vs observed data. Blue shading indicates the invasion time occurred earlier when using the mobility proxy, while red shading indicates a later invasion time when using mobility proxies. We use a relative scale, calculated as the difference between invasion time using mobility proxy and in the baseline (using the empirical mobility data), divided by the largest invasion times across all locations when using the proxy. The seeding location is marked with a red triangle. **E)** Invasion order similarity ($p_n^{first}$). The y-axis shows the proportion of the first $n$ patches invaded in an epidemic model using observed mobility that were also among the first $n$ locations invaded when a mobility proxy was used in the epidemic model. $n$ is the proportion shown on the x-axis. The two lines show different seed locations. The dashed red line shows the expected value if locations are chosen randomly.

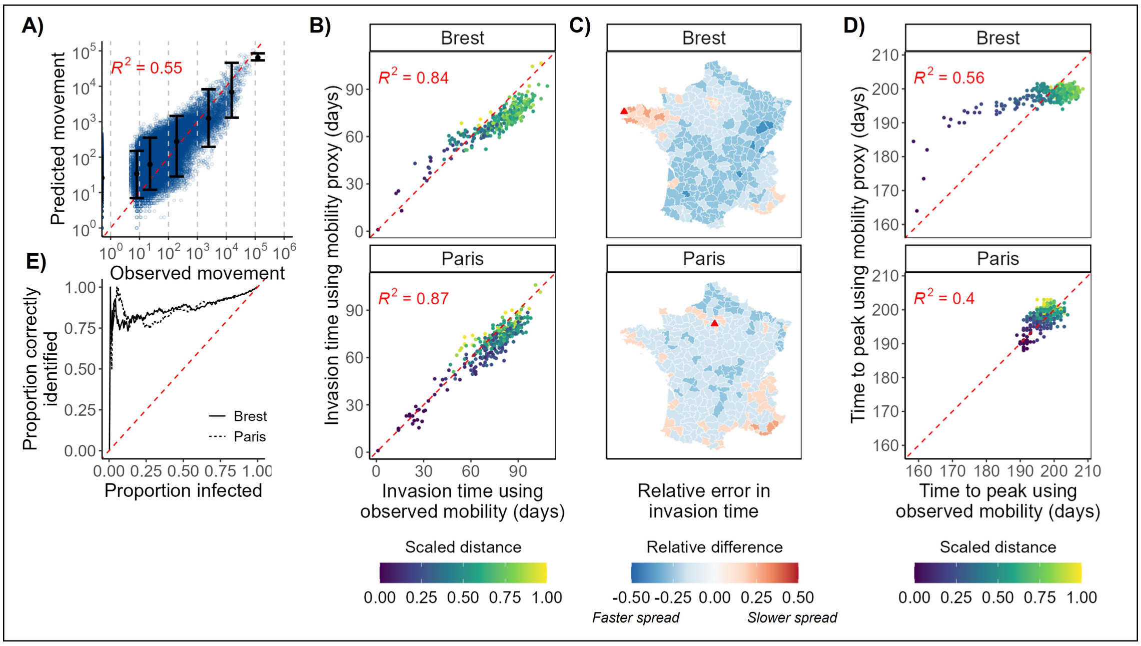


Figure S2: **Mobility predictions and epidemic simulations for France under Suppl. Scenario 2.** In this scenario $p_i^{stay}$ and $p_{ij}^{move}$ are informed by local data (see Table S1 for further scenario details). See caption of Fig. S1 for additional figure details.

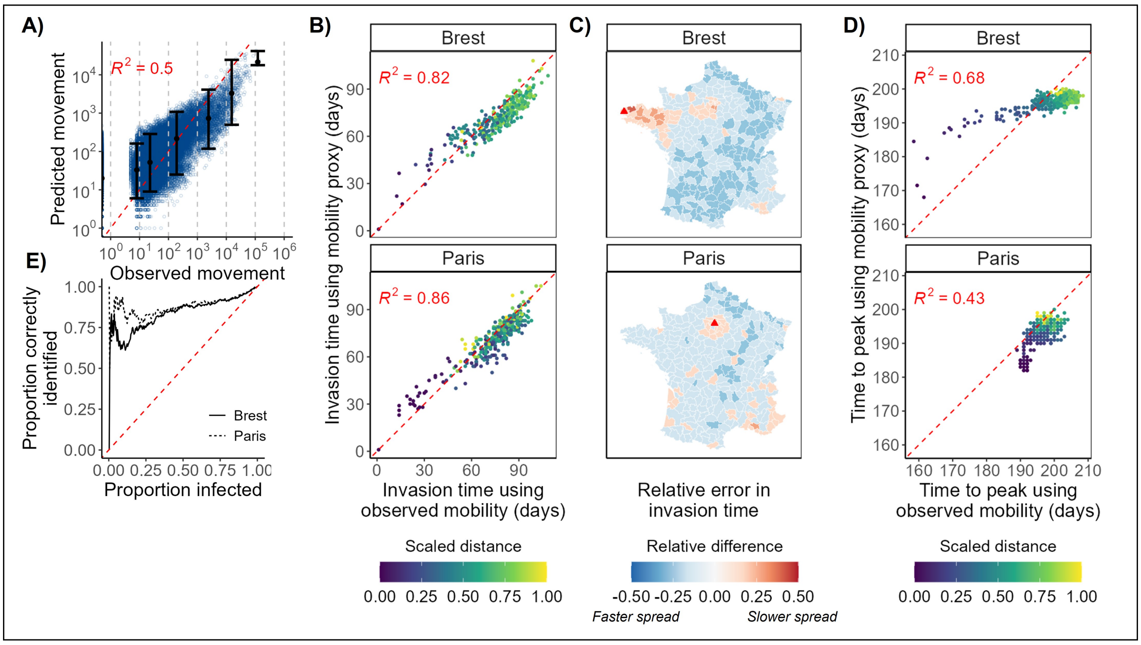


Figure S3: **Mobility predictions and epidemic simulations for France under Suppl. Scenario 3.** In this scenario $p_i^{stay}$ and $p_{ij}^{move}$ are informed by local data but at an aggregated spatial scale (see Table S1 for further scenario details). See caption of Fig. S1 for additional figure details.

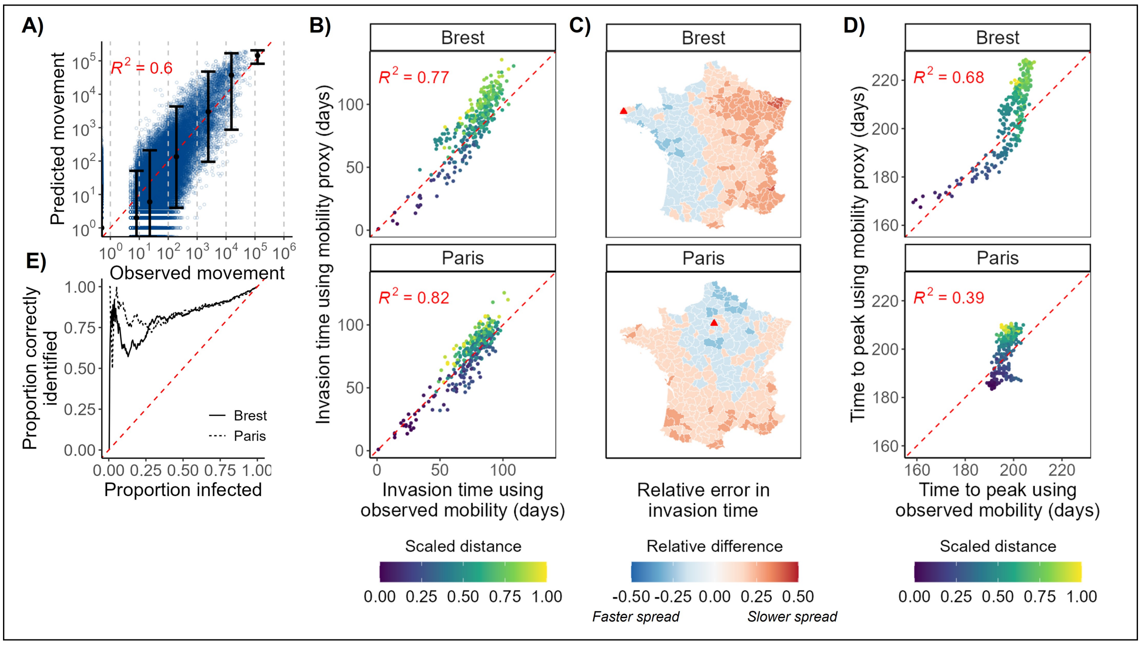


Figure S4: **Mobility predictions and epidemic simulations for France under Suppl. Scenario 4.** In this scenario $p_i^{stay}$ and $p_{ij}^{move}$ are informed by data from Portugal, and a radiation model is used to inform $p_{ij}^{move}$ (see Table S1 for further scenario details). See caption of Fig. S1 for additional figure details.

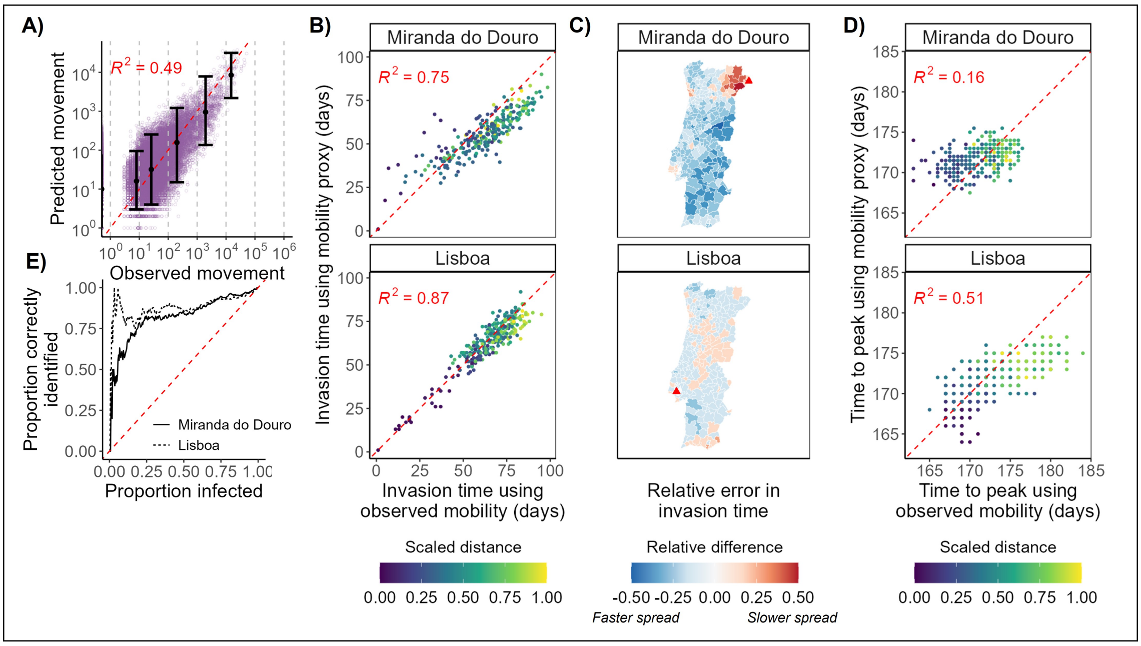


Figure S5: **Mobility predictions and epidemic simulations for Portugal under Suppl. Scenario 1.** In this scenario $p_i^{stay}$ is informed by local data, and $p_{ij}^{move}$ is informed by data from France (see Table S1 for further scenario details). Figure details are as in Fig. S1 except here **A)** shows predicted numbers moving between Portuguese ADM2 units vs observed numbers from mobile phone call records. Each purple dot represents a pair of ADM2 units.

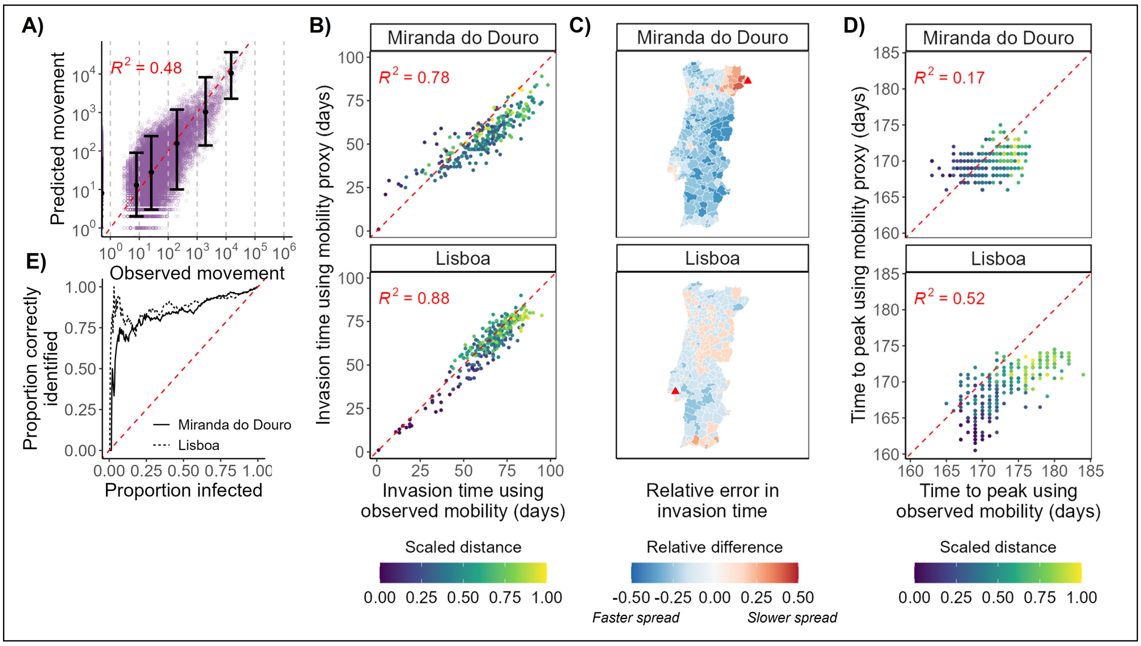


Figure S6: **Mobility predictions and epidemic simulations for Portugal under Suppl. Scenario 2.** In this scenario $p_i^{stay}$ and $p_{ij}^{move}$ are informed by local data (see Table S1 for further scenario details). Figure details are as in Fig. S1 except here **A)** shows predicted numbers moving between Portuguese ADM2 units vs observed numbers from mobile phone call records. Each purple dot represents a pair of ADM2 units.

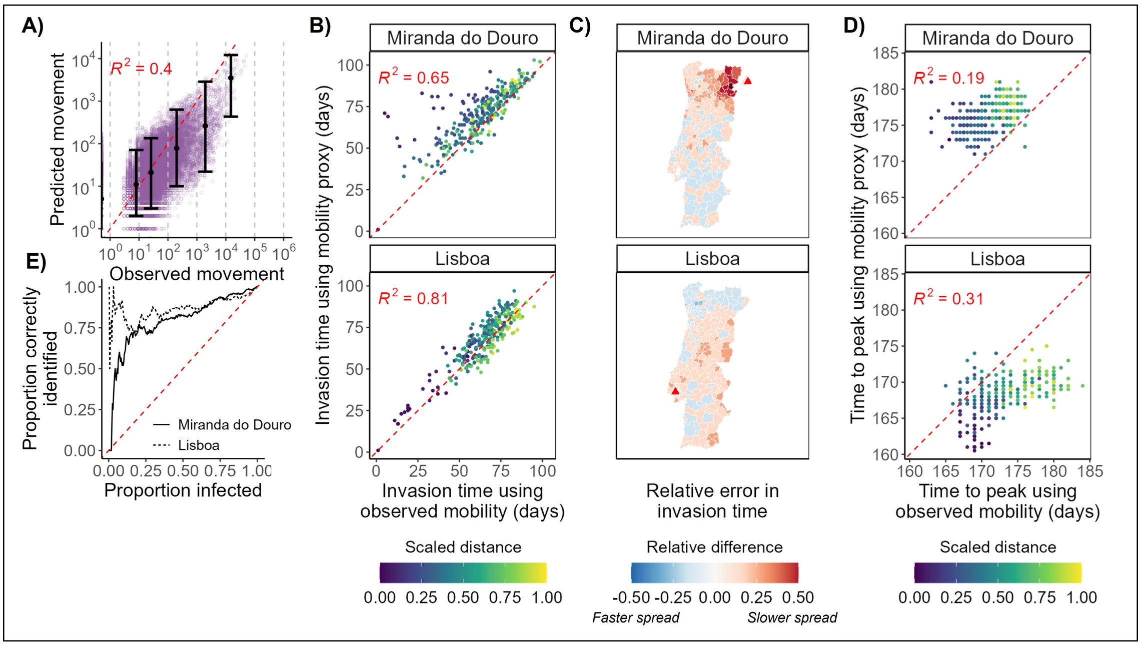


Figure S7: **Mobility predictions and epidemic simulations for Portugal under Suppl. Scenario 3.** In this scenario $p_i^{stay}$ and $p_{ij}^{move}$ are informed by local data but at an aggregated spatial scale (see Table S1 for further scenario details). Figure details are as in Fig. S1 except here **A)** shows predicted numbers moving between Portuguese ADM2 units vs observed numbers from mobile phone call records. Each purple dot represents a pair of ADM2 units.

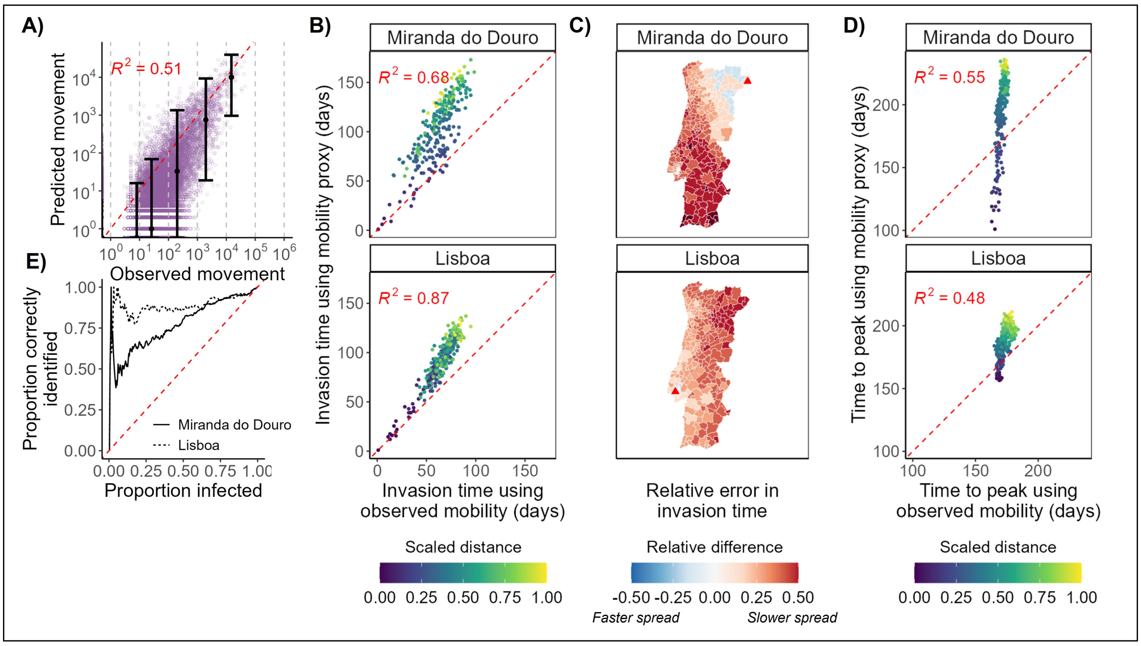


Figure S8: **Mobility predictions and epidemic simulations for Portugal under Suppl. Scenario 4.** In this scenario $p_i^{stay}$ and $p_{ij}^{move}$ are informed by data from France, and a radiation model is used to inform $p_{ij}^{move}$ (see Table S1 for further scenario details). Figure details are as in Fig. S1 except here **A)** shows predicted numbers moving between Portuguese ADM2 units vs observed numbers from mobile phone call records. Each purple dot represents a pair of ADM2 units.

### References


[1] C. T. Lloyd et al. "Global spatio-temporally harmonised datasets for producing high-resolution gridded population distribution datasets". In: *Big Earth Data* 3.2 (2019), pp. 108–139.

[2] WorldPop. "Open Spatial Demographic Data and Research". In: *Available from: https://www.worldpop.org* (2022).

[3] U. of California. "Global Administrative Areas (GADM)". In: *Available from: https://www.worldpop.org* (2022).

[4] R Core Team. *R: A Language and Environment for Statistical Computing.* R Foundation for Statistical Computing. Vienna, Austria, 2021. URL: `https://www.R-project.org/`.

[5] "movement package (R)". In: *https://github.com/SEEG-Oxford/movement* (2022).

[6] F. Simini et al. "A universal model for mobility and migration patterns". In: *Nature* 484.7392 (2012), pp. 96–100. ISSN: 1476-4687. DOI: `10.1038/nature10856`. URL: `https://doi.org/10.1038/nature10856`.

[7] M. Tizzoni et al. "On the Use of Human Mobility Proxies for Modeling Epidemics". In: *PLOS Computational Biology* 10.7 (2014), e1003716. DOI: `10.1371/journal.pcbi.1003716`. URL: `https://doi.org/10.1371/journal.pcbi.1003716`.
